# Trends in known and undiagnosed diabetes, HbA1c levels, cardio-metabolic risk factors and diabetes treatment target achievement in repeated cross-sectional surveys – The Tromsø Study 1994-2016

**DOI:** 10.1101/2020.10.30.20222117

**Authors:** Petja Lyn Langholz, Tom Wilsgaard, Inger Njølstad, Rolf Jorde, Laila Arnesdatter Hopstock

**Affiliations:** Department of Community Medicine, UiT The Arctic University of Norway, Tromsø, Norway; Tromsø Endocrine Research Group, Department of Clinical Medicine, UiT The Arctic University of Norway, Tromsø, Norway; Division of Internal Medicine, University Hospital of North Norway, Tromsø, Norway

## Abstract

**Objectives:** The aim of this study was to investigate time trends in known and undiagnosed diabetes, HbA1c levels, and other cardio-metabolic risk factors in the general population as well as treatment target achievement among those with diabetes.

**Design and setting:** Repeated cross-sectional surveys in the population-based Tromsø Study.

**Methods:** We used age-adjusted generalized estimating equation models to study trends in self-reported and undiagnosed (HbA1c ≥6.5%) diabetes, cardio-metabolic risk factors and the metabolic syndrome in 27281 women and men aged 40-84 years examined in up to four surveys of the Tromsø Study between 1994 and 2016. Further, we analyzed trends in diabetes treatment target achievement.

**Results:** During 1994-2016 diabetes prevalence increased in women (2.3% to 3.9%) and men (2.4% to 5.3%) and in all age-groups, while the proportion of undiagnosed diabetes in women (33% to 20%) and men (37% to 27%) decreased. Blood pressure and total cholesterol decreased, while waist circumference increased in participants with and without diabetes, leading to a relatively stable prevalence of the metabolic syndrome throughout the study period. There was a marginal increase in HbA1c levels among participants without diabetes. Only half of those with diabetes achieved the treatment target of HbA1c ≤7.0%.

**Conclusion:** In the last two decades diabetes prevalence increased, while the proportion of undiagnosed diabetes declined. The prevalence of the metabolic syndrome remained stable throughout, driven by opposing trends with an increase in obesity and a decrease in other cardio-metabolic risk factors. HbA1c treatment target achievement did not improve.

## INTRODUCTION

The prevalence of diabetes is increasing worldwide, partly driven by a growing and older population.[1] In 2019 it was estimated that 9.3% of the world’s adult population have diabetes, which corresponds to 463 million people of whom half are assumed to be unaware of their condition.[2] Patients with diabetes are at high risk of cardiovascular disease (CVD), diseases of the kidneys and eyes, and death.[3] Avoiding hyperglycemia is considered a crucial factor for reducing the risk of these subsequent complications and mortality.[4] Monitoring trends in HbA1c treatment target achievement is therefore important. A World Health Organization report from 2011 concluded that glycated hemoglobin (HbA1c) can be used to diagnose diabetes, with a recommended cut-off value of HbA1c ≥6.5% (i.e. 48 mmol/mol).[5] Thus, HbA1c can be used for diabetes screening in the population.

The metabolic syndrome is a set of cardio-metabolic risk factors for CVD including abdominal obesity, elevated blood pressure, unfavorable blood lipid profile, and impaired fasting glucose or insulin resistance.[6] In 2010, modifiable cardio-metabolic risk factors accounted for 63% of deaths from CVD, chronic kidney disease and diabetes - one in every five deaths worldwide.[7] Over the past decades, a paradoxical trend in cardio-metabolic risk factors has been observed in high-income countries with substantial declines in blood pressure and blood cholesterol, and stagnancy or increase in body mass index (BMI) and fasting glucose.[7] Trends in HbA1c have been studied in patients with diabetes,[8] but trends in HbA1c in the general population are lacking. Thus, exploring trends in cardio-metabolic risk factors in the general population, among individuals both with and without diabetes, is of interest.

The aim of this study was to investigate trends in both known and undiagnosed diabetes, associated cardio-metabolic risk factors including HbA1c, and HbA1c treatment target achievement over two decades in a general population of adult and elderly women and men.

## METHODS

### Sample

The Tromsø Study[9,10] is a population-based study conducted by UiT The Arctic University of Norway in the municipality of Tromsø, Northern Norway, consisting of seven surveys conducted to date; Tromsø 1 (1974), Tromsø 2 (1979–1980), Tromsø 3 (1986–1987), Tromsø 4 (1994–1995), Tromsø 5 (2001), Tromsø 6 (2007–2008) and Tromsø 7 (2015–2016). Total birth cohorts and random samples of the inhabitants were invited (participation 65-79%), and 45473 women and men have participated in one or more surveys. Data collection consisted of questionnaires, biological sampling and clinical examinations.

Eligible for the present analysis were 32611 participants aged 40 to 84 years, who attended at least one of the four surveys conducted between 1994 and 2016: Tromsø 4: first visit: n=16857, second visit: n=7428; Tromsø 5: n=7415; Tromsø 6: n=12367; Tromsø 7: n=20841. 51% of participants attended one survey, 27% two, 15% three and 7% attended all four surveys, amounting to 57480 observations in total. For analysis of self-reported diabetes, we excluded participants who did not consent to research (182 observations), pregnant women (69 observations) and participants with missing data on self-reported diabetes (1049 observations), leaving 32171 participants (56180 observations) for analysis. In Tromsø 4 HbA1c was only measured in the second visit, so 28262 participants (46904 observations) remained available for analysis of undiagnosed diabetes, metabolic syndrome and diabetes treatment target achievement. In this set of analyses we further excluded participants with missing data on HbA1c (2495 observations) and cardio-metabolic risk factors (1438 observations). Altogether, 27281 participants (42971 observations) were included in the analyses.

The Regional Committee of Medical and Health Research Ethics and the Norwegian Data Protection Authority have approved the Tromsø Study, and all procedures were carried out in accordance with the 1964 Helsinki declaration and its later amendments. The participants gave written informed consent.

### Data collection and laboratory analyses

Data collection was performed by standard methods and trained personnel. Non-fasting blood samples were drawn from an antecubital vein after a brief stasis with the participant seated. All laboratory analyses were performed at the University Hospital of Northern Norway (UNN), except for HbA1c in Tromsø 4 (performed at the Laboratory for Metabolic Research at UiT) and Tromsø 5 (performed at the study site laboratory in accordance with the hospital gold standard).

HbA1c was analyzed by immunoturbidimetry with Cobras Mira Plus (Unimate 5 HbA1c, F. Hoffmann-La Roche AG, Basel, Switzerland) (Tromsø 4), DCA 2000 (Bayer Diagnostics, Tarrytown, NY, USA) (Tromsø 5), and by high performance liquid chromatography (HPLC) with Variant II (Bio-Rad Laboratories Inc., Hercules, CA, USA) (Tromsø 6) and Tosoh G8 (Tosoh Bioscience, San Francisco, USA) (Tromsø 7).

Serum total cholesterol, high-density lipoprotein (HDL) cholesterol and triglycerides were analyzed by enzymatic colorimetric methods with Hitachi 737 (Boehringer-Mannheim, Mannheim, Germany) (Tromsø 4), Hitachi 917 (Tromsø 5), Modular PPE (Tromsø 6) and Cobas 8000 c702 (Tromsø 7) (all: Roche Diagnostics, Mannheim, Germany). HDL cholesterol was measured after precipitation of lower-density lipoprotein with heparin and manganese chloride (Tromsø 4) or directly (Tromsø 5-7).

Waist circumference was measured with a measuring tape at the umbilical level. Blood pressure was measured three times with one-minute intervals on the participant’s right upper arm with a properly sized cuff based on arm circumference after two minutes seated rest using an oscillometric digital automatic device (Tromsø 4-5: Dinamap Vital Signs Monitor; Critikon, USA; Tromsø 6–7: Dinamap ProCare 300 monitor, GE Healthcare, Norway). The mean of the last two recordings was used in the analyses.

### Diabetes definition

Self-reported diabetes was assessed via questionnaires: *“Do you have or have you had diabetes?”* to which those who answered *“Yes, now”* were defined as having diabetes. Participants who answered *“Yes, previously”* or *“No, never”* with HbA1c levels ≥6.5% (48 mmol/mol) were defined as having undiagnosed diabetes (unawareness). We cannot distinguish between type 1 and type 2 diabetes, but it is expected that the vast majority of cases are type 2 diabetes. Excluding participants with self-reported age at diabetes diagnosis below 20 years did not alter the results.

### Metabolic syndrome definition

The metabolic syndrome was defined using a modified version of the Adult Treatment Panel (ATP) III definition from 2001, in which contrary to other definitions evidence of insulin resistance is not a requirement.[6,11] ATP III includes five risk factors of which at least three have to be present to identify the metabolic syndrome. As a proxy for elevated fasting glucose concentration we used self-reported diabetes or HbA1c ≥ 6.5%. The other four criteria were in accordance with ATP III defined as follows: abdominal obesity (waist circumference ≥102 cm for men and ≥88 cm for women), low HDL cholesterol (<1.03 mmol/L for men and <1.29 mmol/L for women), elevated triglycerides (≥1.7 mmol/L) and hypertension (blood pressure ≥130/85 mmHg or use of antihypertensive medication).

### Medication use and HbA1c treatment target definition

Self-reported use of medication, antihypertensives and antidiabetics (insulin and/or diabetes tablets), was obtained via questionnaires, except in participants younger than 70 years in Tromsø 5, where questions about insulin use were not included. To define achievement of treatment target among participants with diabetes throughout the surveys, the following HbA1c thresholds from the concurrent national diabetes guidelines were used; Tromsø 4: HbA1c <7.5% (>70 years: HbA1c <8.5%);[12] Tromsø 5: HbA1c <7.5% (>75 years: HbA1c <9.0%);[13,14] Tromsø 6: HbA1c <7.5% (>75 years: HbA1c <9.0%)[14] and Tromsø 7: HbA1c ≤7.0%.[15]

### Participant feedback

Standard letters with feedback on selected measurements were sent to all participants after participation. Additional feedback was given to participants (by telephone or letter dependent on severity) above specific cut-off values. For glucose, the following cut-off values were used: Tromsø 4 (second visit): glucose >8.0 mmol/L (special letter); Tromsø 5: no self-reported diabetes and glucose ≥11.0 mmol/L (standard letter), ≥16 mmol/L (special letter), ≥25.0 mmol/L (telephone); Tromsø 6: glucose ≥9.0 mmol/L (standard letter), no self-reported diabetes and glucose >11.0 mmol/L (special letter), >20.0 mmol/L (telephone); Tromsø 7: no self-reported diabetes and glucose >20.0 mmol/L (telephone). For HbA1c the following cut-off values were used: Tromsø 6: HbA1c ≥7.0% and no self-reported diabetes (standard letter); Tromsø 7: HbA1c ≥6.5% (standard letter). It was up to the participants to decide whether to consult their general practitioner or other health care providers about their examination results.

### Statistics

Descriptive characteristics are presented as means with standard deviation (SD) for continuous variables and proportions and frequency for binary variables. Generalized estimating equation (GEE) models were used for age-adjustment of mean values and proportions, and to test for age-adjusted linear trends using year of survey as a continuous time variable. Odds ratios (OR) and linear regression coefficients were reported as effect sizes for linear trend in binary and continuous variables, respectively. The unit for the regression coefficient and OR was reported per 10 years. An exchangeable within-group correlation structure was specified to control for dependencies between repeated measures within individuals across the four surveys. In a sensitivity analysis we excluded all observations which followed feedback about elevated glucose or HbA1c levels in a previous Tromsø Study survey. All analyses were performed using Stata version 16.0 (StataCorp LLP, College Station, TX, USA).

## RESULTS

During the study period from 1994 to 2016 there was a linear increase in prevalence of self-reported diabetes in both sexes and all age-groups; from 2.3% to 3.9% in women, and 2.4% to 5.3% in men (Table 1). The proportion of undiagnosed diabetes among participants with diabetes (i.e. unawareness) decreased during the study period to a level of 20% in women and 27% for men in 2016 (Table 2). These results were not significantly altered after excluding participants who had received feedback about elevated glucose or HbA1c levels in a previous survey (Supplementary Table 1). There was a marginal increase in mean HbA1c levels among participants without diabetes (Table 3).

**Table 1.** Trends in age-adjusted prevalence of self-reported diabetes among women and men aged 40-84. The Tromsø Study 1994-2016.

|  | Tromsø 4 | Tromsø 5 | Tromsø 6 | Tromsø 7 | OR (95% CI)* | P value |
| --- | --- | --- | --- | --- | --- | --- |
| Prevalence, % (n) |  |  |  |  |  |  |
| **Women** | N=8653 | N=4031 | N=6394 | N=10551 |  |  |
| Total | 2.3 (227) | 2.7 (157) | 3.9 (295) | 3.9 (445) | 1.31 (1.23, 1.40) | < 0.001 |
| 40-49 years | 0.7 (22) | 1.1 (8) | 2.0 (40) | 2.2 (71) | 1.70 (1.39, 2.09) | < 0.001 |
| 50-59 years | 1.3 (29) | 1.9 (14) | 3.3 (44) | 3.1 (98) | 1.46 (1.21, 1.75) | < 0.001 |
| 60-69 years | 3.5 (58) | 3.8 (58) | 5.4 (109) | 5.6 (141) | 1.27 (1.11, 1.45) | 0.001 |
| 70-84 years | 7.4 (118) | 7.1 (77) | 8.9 (102) | 9.0 (135) | 1.12 (1.00, 1.26) | 0.060 |
| **Men** | N=8020 | N=3161 | N=5694 | N=9676 |  |  |
| Total | 2.4 (201) | 3.2 (142) | 5.2 (325) | 5.3 (535) | 1.45 (1.36, 1.55) | < 0.001 |
| 40-49 years† | 1.1 (37) | 1.7 (10) | 2.4 (39) | 2.5 (77) | 1.47 (1.23, 1.75) | < 0.001 |
| 50-59 years | 1.9 (42) | 3.7 (14) | 4.8 (55) | 4.7 (128) | 1.48 (1.27, 1.73) | < 0.001 |
| 60-69 years | 3.9 (57) | 4.8 (60) | 7.9 (151) | 7.5 (188) | 1.36 (1.20, 1.54) | < 0.001 |
| 70-84 years | 5.8 (65) | 6.5 (58) | 9.1 (80) | 9.7 (142) | 1.31 (1.14, 1.50) | < 0.001 |

Proportions, odds ratios (OR) with confidence intervals (CI) and p-value for trend are adjusted for age across surveys using generalized estimating equation (GEE) models. N represents crude numbers.
\*Odds ratios for proportions are presented per 10 years.
†Logistic regression used for age-adjustment (GEE model did not converge).

**Table 2.** Age-adjusted prevalence of undiagnosed diabetes among women and men with diabetes aged 40-84. The Tromsø Study 1994-2016.

|  | Tromsø 4 | Tromsø 5 | Tromsø 6 | Tromsø 7 | OR (95% CI)* | P value |
| --- | --- | --- | --- | --- | --- | --- |
| Diabetes prevalence, % (n) |  |  |  |  |  |  |
| **Women** | N=120 | N=244 | N=388 | N=540 |  |  |
| Undiagnosed† | 32.6 (32) | 42.4 (96) | 32.1 (117) | 20.4 (111) | 0.62 (0.52, 0.73) | <0.001 |
| **Men** | N=132 | N=242 | N=440 | N=715 |  |  |
| Undiagnosed† | 36.5 (38) | 51.4 (121) | 35.1 (142) | 27.4 (193) | 0.67 (0.57, 0.78) | <0.001 |

Proportions, odds ratios (OR) with confidence intervals (CI) and p-value for trend are adjusted for age across surveys using generalized estimating equation (GEE) models. N represents crude numbers.
\*Odds ratios for proportions are presented per 10 years.
†No self-reported diabetes and HbA1c $\geq 6.5\%$ .

**Table 3.**
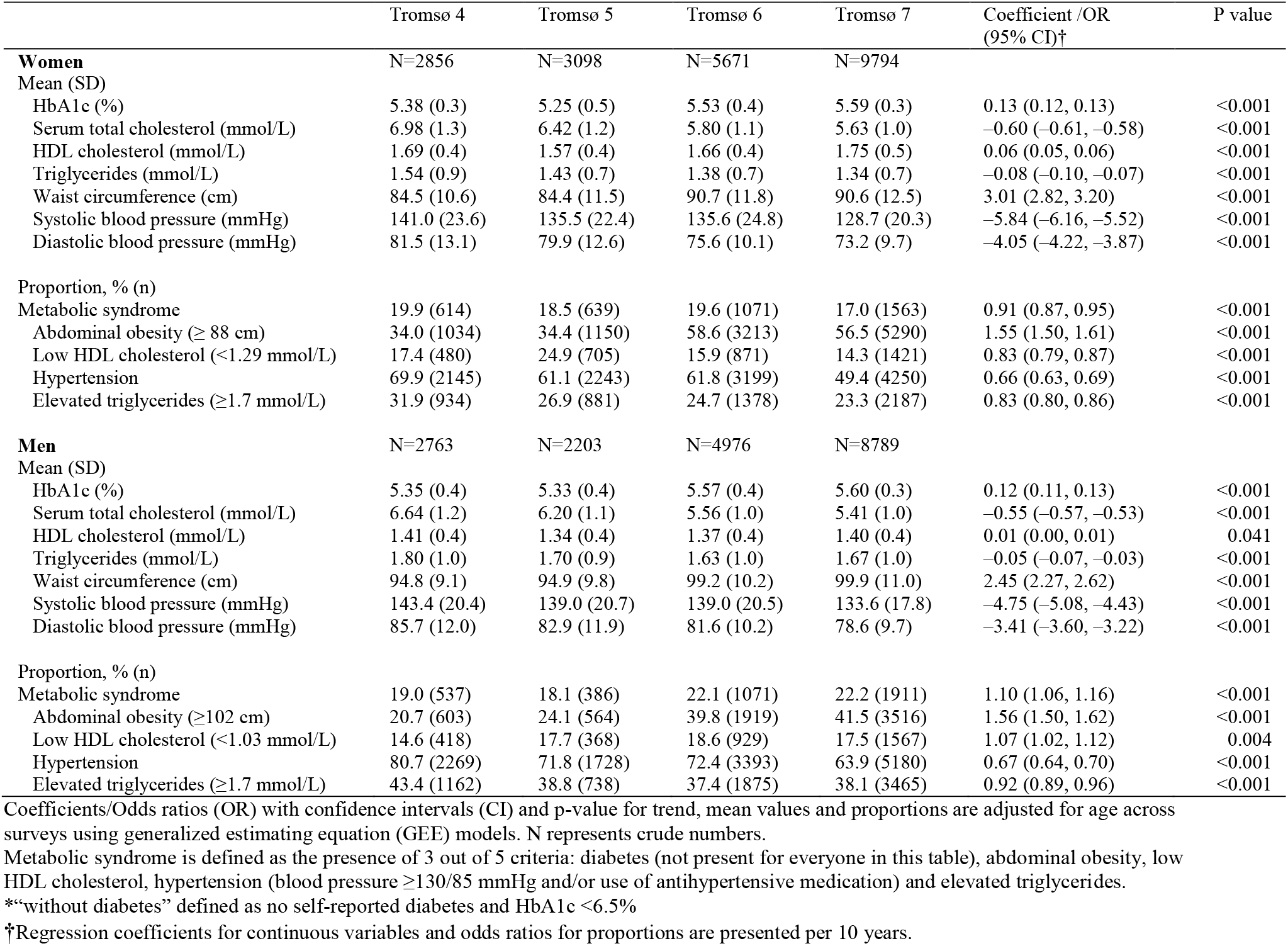
Age-adjusted mean values and trend in cardio-metabolic risk factors and prevalence of metabolic syndrome among women and men without diabetes* aged 40-84. The Tromsø Study 1994-2016.

During the study period, abdominal obesity increased substantially both in women and men with and without diabetes, while total cholesterol, triglycerides and blood pressure decreased (Table 3, 4). Thus, although driven by changing risk factors levels, the prevalence of the metabolic syndrome was fairly stable throughout the study period, in both women (78-85% and 17-20%) and men (73-78% and 18-22%) with and without diabetes, respectively. In the total study sample, the prevalence of metabolic syndrome varied between 22% and 25% from 1994 to 2016. The proportion of HbA1c treatment target achievement according to the respective concurrent guidelines decreased during the study period in both women and men with diabetes (Table 5). Stratified by medication use, the decreasing trend was larger among those reporting use of antidiabetics. In the latest survey, the treatment target was reached by 43.2% female and 38.4% male users of antidiabetics compared to 76.5% and 70.5% female and male non-users, respectively. When applying the same treatment target (HbA1c ≤7.0% or <7.5%) to all surveys, the variation in target achievement across surveys was small.

**Table 4.** Age-adjusted mean values and trend in cardio-metabolic risk factors and prevalence of metabolic syndrome among women and men with diabetes* aged 40-84. The Tromsø Study 1994-2016.

|  | Tromsø 4 | Tromsø 5 | Tromsø 6 | Tromsø 7 | Coefficient /OR<br>(95% CI) † | P value |
| --- | --- | --- | --- | --- | --- | --- |
| **Women** | N=120 | N=244 | N=388 | N=540 |  |  |
| Mean (SD) |  |  |  |  |  |  |
| HbA1c (%) | 7.38 (2.2) | 7.19 (1.5) | 7.05 (1.2) | 7.23 (1.2) | -0.01 (-0.12, 0.10) | 0.871 |
| Serum total cholesterol (mmol/L) | 6.89 (1.2) | 6.22 (1.2) | 5.48 (1.2) | 5.31 (1.1) | -0.68 (-0.77, -0.59) | <0.001 |
| HDL cholesterol (mmol/L) | 1.48 (0.4) | 1.35 (0.4) | 1.37 (0.4) | 1.46 (0.5) | 0.04 (0.01, 0.06) | 0.007 |
| Triglycerides (mmol/L) | 2.19 (1.1) | 2.07 (1.1) | 1.94 (1.0) | 1.99 (1.1) | -0.07 (-0.15, 0.01) | 0.110 |
| Waist circumference (cm) | 95.8 (12.2) | 94.8 (13.3) | 100.3 (13.3) | 101.4 (14.8) | 3.30 (2.37, 4.22) | <0.001 |
| Systolic blood pressure (mmHg) | 158.6 (26.8) | 147.7 (22.0) | 145.0 (24.2) | 138.0 (20.9) | -8.67 (-10.34, -7.00) | <0.001 |
| Diastolic blood pressure (mmHg) | 85.6 (16.3) | 81.1 (13.2) | 75.7 (9.9) | 73.8 (9.4) | -5.20 (-6.03, -4.36) | <0.001 |
| Proportion, % (n) |  |  |  |  |  |  |
| Metabolic syndrome | 84.6 (103) | 78.4 (191) | 84.2 (328) | 83.2 (448) | 1.06 (0.87, 1.28) | 0.572 |
| Abdominal obesity (≥88 cm) | 74.0 (88) | 73.3 (170) | 81.5 (318) | 83.6 (458) | 1.40 (1.17, 1.66) | 0.001 |
| Low HDL cholesterol (<1.29 mmol/L) | 35.6 (45) | 45.2 (108) | 43.6 (170) | 40.0 (217) | 0.98 (0.84, 1.13) | 0.740 |
| Hypertension | 89.5 (109) | 86.7 (216) | 87.1 (331) | 82.6 (422) | 0.75 (0.59, 0.95) | 0.018 |
| Elevated triglycerides (≥1.7 mmol/L) | 64.8 (79) | 53.3 (131) | 52.1 (201) | 54.5 (295) | 0.92 (0.79, 1.07) | 0.272 |
| **Men** | N=132 | N=242 | N=440 | N=715 |  |  |
| Mean (SD) |  |  |  |  |  |  |
| HbA1c (%) | 7.29 (1.6) | 7.35 (1.3) | 7.35 (1.4) | 7.42 (1.3) | 0.06 (-0.04, 0.16) | 0.226 |
| Serum total cholesterol (mmol/L) | 6.57 (1.3) | 5.88 (1.3) | 5.06 (1.1) | 4.84 (1.2) | -0.73 (-0.81, -0.64) | <0.001 |
| HDL cholesterol (mmol/L) | 1.27 (0.4) | 1.20 (0.3) | 1.21 (0.3) | 1.24 (0.4) | 0.00 (-0.02, 0.03) | 0.799 |
| Triglycerides (mmol/L) | 2.38 (1.5) | 2.18 (1.4) | 2.03 (1.3) | 2.14 (1.7) | -0.05 (-0.16, 0.05) | 0.339 |
| Waist circumference (cm) | 101.4 (11.6) | 102.6 (11.9) | 106.1 (11.5) | 107.3 (12.3) | 2.80 (2.00, 3.60) | <0.001 |
| Systolic blood pressure (mmHg) | 153.2 (19.8) | 145.0 (19.2) | 143.6 (20.8) | 137.9 (17.9) | -6.55 (-7.89, -5.21) | <0.001 |
| Diastolic blood pressure (mmHg) | 85.8 (13.3) | 83.7 (12.0) | 80.6 (10.4) | 77.2 (9.8) | -4.33 (-5.07, -3.58) | <0.001 |
| Proportion, % (n) |  |  |  |  |  |  |
| Metabolic syndrome | 77.8 (102) | 72.5 (175) | 77.3 (338) | 77.7 (559) | 1.07 (0.91, 1.27) | 0.420 |
| Abdominal obesity (≥102 cm) | 46.2 (57) | 54.7 (126) | 65.0 (282) | 67.2 (488) | 1.44 (1.25, 1.66) | <0.001 |
| Low HDL cholesterol (<1.03 mmol/L) | 28.9 (41) | 30.3 (74) | 35.5 (155) | 37.4 (273) | 1.20 (1.04, 1.39) | 0.013 |
| Hypertension | 95.5 (126) | 87.5 (215) | 88.5 (385) | 84.5 (590) | 0.66 (0.52, 0.84) | 0.001 |
| Elevated triglycerides (≥1.7 mmol/L) | 62.1 (82) | 54.7 (129) | 52.4 (227) | 55.5 (403) | 0.96 (0.83, 1.11) | 0.564 |

Coefficients/Odds ratios (OR) with confidence intervals (CI) and p-value for trend, mean values and proportions are adjusted for age across surveys using generalized estimating equation (GEE) models. N represents crude numbers.
Metabolic syndrome defined as the presence of 3 out of 5 criteria: diabetes (present for everyone in this table), abdominal obesity, low HDL cholesterol, hypertension (blood pressure ≥130/85 mmHg and/or use of antihypertensive medication) and elevated triglycerides.
\*Diabetes defined as self-reported diabetes or HbA1c ≥6.5%.
†Regression coefficients for continuous variables and odds ratios for proportions are presented per 10 years.

**Table 5.**
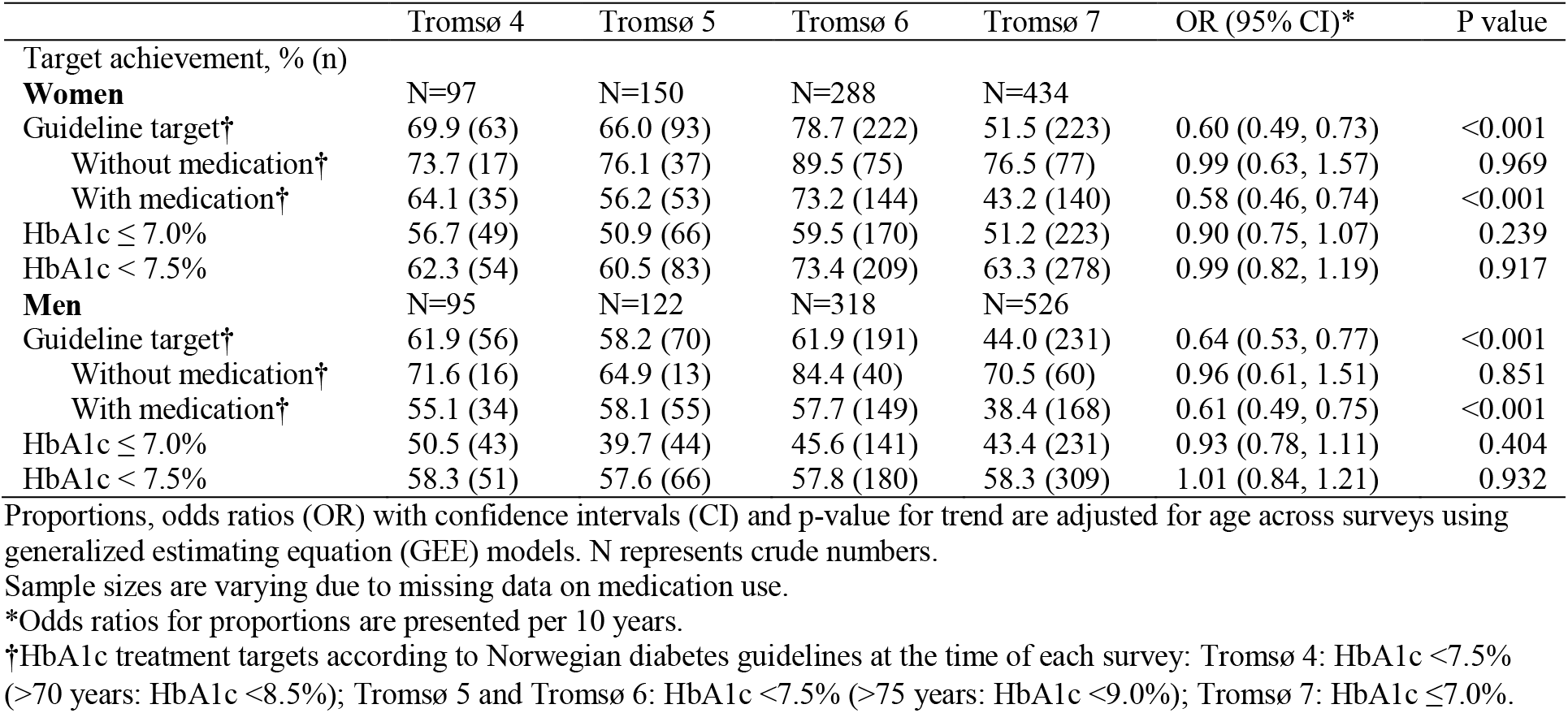
Age-adjusted prevalence and trend in HbA1c treatment target achievement among women and men with self-reported diabetes aged 40-84. The Tromsø Study 1994-2016.

## DISCUSSION

We found an increase in diabetes prevalence between 1994 and 2016 in both sexes and all age groups, while the proportion of undiagnosed diabetes declined. All cardio-metabolic risk factors decreased, except for HbA1c which increased marginally, and abdominal obesity which showed a substantial increase. In total, the prevalence of metabolic syndrome was relatively stable. Further, the proportion of participants with diabetes meeting the HbA1c treatment target did not improve throughout the study period.

### Diabetes prevalence

The observed increase in diabetes prevalence in our study is consistent with findings from national register-based studies from Norway,[16] Sweden[17] and Denmark,[18] although the increase seems to be flattening towards the end of the study period. A study based on data from the Norwegian Prescription Database, the Norwegian Patient Registry and the primary care database found that between 2009 and 2014 the prevalence of type 2 diabetes in Norway increased from 4.9% to 6.1% in the age group 30–89.[16] Similarly, a study based on the Swedish Prescribed Drug Register found an increase of diabetes prevalence in the Swedish population from 5.8 to 6.8% between 2007 and 2013.[17] In accordance with previous studies,[16-18] diabetes prevalence in our study was higher among men than women. As shown in register-based studies,[16,17] increase of diabetes prevalence does not seem to be accompanied by a rise in diabetes incidence. Thus, the increased prevalence could partially be explained by earlier diagnosis and increased life expectancy, as previously suggested.[16]

### Undiagnosed diabetes

The Diabetes Atlas from 2019 by the International Diabetes Federation reported an estimated prevalence of undiagnosed diabetes in Europe of 40%,[2] which is considerably higher than our findings. Further, we found a decrease in the proportion of undiagnosed diabetes during the study period. The observed prevalence by the end of the study period corresponds to findings from a Danish study from 2011,[19] which estimated that 24% of all diabetes cases in Denmark remained undiagnosed, based on data from Danish health registers and population-studies using HbA1c and self-reported diabetes. HbA1c was introduced as the primary diagnostic tool in Norway in 2012, which could have increased population awareness and health care screening, and thus explain the distinct decline of undiagnosed diabetes between the sixth (2007-2008) and seventh (2015-2016) survey of the Tromsø Study.

### Metabolic risk factors and the metabolic syndrome

We found a considerable increase in mean waist circumference and prevalence of abdominal obesity in participants both with and without diabetes. At the same time, we found that other cardio-metabolic risk factors like total cholesterol and blood pressure decreased. This paradoxical trend is in accordance with findings from high-income countries globally,[7] in the Norwegian population-based HUNT study[20,21] and in previous publications from the Tromsø Study of longitudinal trends using repeated measures in the same individuals.[22-24] This also corresponds to findings from the National Health and Nutrition Examination Surveys in the United States 1999-2004, where 40% of obese individuals were considered metabolically healthy according to the ATP III definition.[25]

The overall prevalence of the metabolic syndrome among participants both with and without diabetes remained relatively stable throughout the study period. This seems to a large extent to be driven by the simultaneous trends of increase in abdominal obesity and decrease in hypertension. Mean HbA1c levels among participants without diabetes were relatively stable throughout. As expected, both mean levels and proportions of elevated single risk factors, as well as the proportion of metabolic syndrome, were considerably higher among participants with diabetes compared to those without. The overall prevalence of the metabolic syndrome in our study (22-25%) is consistent with findings from a study of cohorts from 10 European countries and the United States using the ATP III criteria, which reported a total prevalence of metabolic syndrome of 24%.[26]

### Treatment target achievement

We found no improvement in achievement of guideline-based HbA1c targets for diabetes treatment contrary to previous findings.[27,28] A Norwegian study of diabetes care in general practice found an improvement in HbA1c target achievement in 2014 compared to 2005.[28] However, similarly to our study the authors found that target achievement was lower among patients using antidiabetics compared to non-users. This could be explained by less severe disease among non-users (i.e. diet-regulated diabetes).

### Strengths and limitations

The observed prevalence of diabetes in this population-based sample was lower compared to national register-based studies, which could be due to selection bias towards healthy participants. Further, the feedback given to participants with elevated glucose or HbA1c levels may have led to a diabetes diagnosis from a health care provider. Yet, excluding observations that followed feedback did not alter the conclusion of a decreasing prevalence of undiagnosed diabetes. Another limitation is the use of a modified version of the ATP III criteria for the metabolic syndrome, including the definition of elevated triglycerides based on non-fasting samples. However, a previous publication from the Tromsø Study about the metabolic syndrome and risk factors for subsequent diabetes found that using non-fasting triglycerides did not alter the results significantly.[29] Another limitation of this study is the lack of possibility to distinguish between diabetes type 1 and type 2. A major strength of this study compared to register-based studies is the availability of HbA1c data and thus the possibility to investigate trends in both known and undiagnosed diabetes. Another strength is the long observation time over more than two decades, the inclusion of a large population-sample of both adults and elderly, and moreover, the possibility to study coherent trends in several cardio-metabolic risk factors as well as treatment target achievement.

### Conclusion

In this population-based study examining trends in diabetes and other cardio-metabolic risk factors in the years 1994 to 2016, we found an increase in diabetes prevalence in both sexes and all age groups, but a decline in undiagnosed diabetes. The prevalence of the metabolic syndrome was relatively stable throughout the study period, with a shift from elevated blood pressure and an unfavorable blood lipid profile to increased waist circumference. The observed lack of improvement in the proportion of patients meeting the HbA1c treatment target should be investigated further.

## Supporting information

STROBE checklist

## Data Availability

The data referred to in the manuscript were used under license for the current study, and so are not publicly available. The data are available upon application to the Tromso Study.

## Contributors

LAH and IN designed the study. RJ and IN contributed to data collection. PLL and TW performed the statistical analysis. All authors contributed to the interpretation of the results. PLL and LAH drafted the manuscript. All authors critically revised the manuscript. All authors read and approved the final manuscript.

## Data sharing

No additional data available.

