## Supplementary Table 1 for "Trends in known and undiagnosed diabetes, HbA1c levels, cardio-metabolic risk factors and diabetes treatment target achievement in repeated cross-sectional surveys – The Tromsø Study 1994-2016"

**Supplementary Table 1.** Age-adjusted prevalence of undiagnosed diabetes among women and men with diabetes aged 40-84, excluding observations following feedback*. The Tromsø Study 1994-2016.

|  | Tromsø 4 | Tromsø 5 | Tromsø 6 | Tromsø 7 | OR (95% CI)† | P value |
| --- | --- | --- | --- | --- | --- | --- |
| Diabetes prevalence, % (n) |  |  |  |  |  |  |
| **Women** | N=120 | N=233 | N=376 | N=505 |  |  |
| Undiagnosed§ | 34.1 (32) | 44.5 (95) | 33.2 (117) | 22.5 (109) | 0.64 (0.54, 0.76) | <0.001 |
| **Men** | N=132 | N=235 | N=428 | N=669 |  |  |
| Undiagnosed§ | 39.3 (38) | 51.7 (118) | 35.6 (140) | 29.4 (183) | 0.69 (0.59, 0.80) | <0.001 |

Proportions, odds ratios (OR) with confidence intervals (CI) and p-value for trend are adjusted for age across surveys using generalized estimating equation (GEE) models. N represents crude numbers.

*All observations of participants who had received feedback about elevated glucose or HbA1c levels in a previous Tromsø Study survey were excluded.

†Odds ratios for proportions are presented per 10 years.

§No self-reported diabetes and HbA1c ≥6.5%.
